# Standard operating procedures for SARS-CoV-2 detection by a clinical diagnostic RT-LAMP assay

**DOI:** 10.1101/2020.06.29.20142430

**Authors:** On behalf of the Crick COVID-19 Consortium, Michael D. Buck, Enzo Z. Poirier, Ana Cardoso, Bruno Frederico, Johnathan Canton, Sam Barrell, Rupert Beale, Richard Byrne, Simon Caidan, Margaret Crawford, Laura Cubitt, Steve Gamblin, Sonia Gandhi, Robert Goldstone, Paul R. Grant, Kiran Gulati, Steve Hindmarsh, Michael Howell, Michael Hubank, Rachael Instrell, Ming Jiang, George Kassiotis, Wei-Ting Lu, James I. MacRae, Iana Martini, Davin Miller, David Moore, Eleni Nastouli, Jerome Nicod, Luke Nightingale, Jessica Olsen, Amin Oomatia, Nicola O’Reilly, Anett Rideg, Ok-Ryul Song, Amy Strange, Charles Swanton, Samra Turajlic, Philip A. Walker, Mary Wu, Caetano Reis e Sousa

## Abstract

The ongoing pandemic of SARS-CoV-2 calls for rapid and cost-effective methods to accurately identify infected individuals. The vast majority of patient samples is assessed for viral RNA presence by RT-qPCR. Our biomedical research institute, in collaboration between partner hospitals and an accredited clinical diagnostic laboratory, established a diagnostic testing pipeline that has reported on more than 40,000 RT-qPCR results since its commencement at the beginning of April 2020. However, due to ongoing demand and competition for critical resources, alternative testing strategies were sought. In this work, we present a clinically-validated standard operating procedure (SOP) for high-throughput SARS-CoV-2 detection by RT-LAMP in 25 minutes that is robust, reliable, repeatable, sensitive, specific, and inexpensive.

## Introduction

The current pandemic caused by novel coronavirus SARS-CoV-2, first detected in late 2019 in the province of Wuhan, China, has rapidly spread worldwide, infecting more than 10 million individuals as of 29 June 2020^1–3^. Infection with SARS-CoV-2 can lead to development of COVID-19, a disease associated with severe acute respiratory syndrome, responsible for hundreds of thousands of deaths globally. Controlling the spread of SARS-CoV-2 relies on the ability of healthcare systems to quickly identify infected individuals, which has mainly relied on RT-qPCR for viral RNA detection^4^. International competition for commercial kits and reagents has negatively impacted the ability of many countries to scale up testing capacity to deal with the increased demand caused by rampant infection. Implementing RT-qPCR testing programs requires specialised laboratory equipment and reagents, presenting additional challenges.

In an effort to increase the diagnostic capacity for SARS-CoV-2 infection in the UK, the Francis Crick Institute, a biomedical research institute based in London, rapidly repurposed its staff and facilities in late March 2020 to serve as a clinical diagnostic testing facility through a partnership between a major local healthcare provider (University College London Hospitals National Health Services Trust) and an accredited clinical diagnostic laboratory (Health Services Laboratories, HSL), termed the CRICK COVID-19 Consortium (CCC)^5^. The pipeline utilises a series of in-house buffers to first inactivate patient samples received from care homes and hospitals, and to then extract RNA before using a CE marked commercial kit to detect SARS-CoV-2 by RT-qPCR. Patient results are reported through a custom online web portal that interfaces with the reference laboratory^5^. In order to avoid dependence on any singular testing methodology, to continue increasing testing capacity, and to provide a potential means to deliver diagnostics at the point-of-care, the CCC was also tasked with developing and validating alternative SARS-CoV-2 testing strategies.

Herein, we describe the use of loop mediated isothermal amplification PCR coupled with reverse transcription (RT-LAMP) as a robust method for SARS-CoV-2 detection in clinical specimens^6^. The entire strand displacement and amplification procedure is carried out at a single temperature in less than 30 minutes, alleviating the need for a thermocycler qPCR system. Diagnostic tests utilising this technique have been developed for RNA viruses and other pathogens^7–9^, and there are now several preprints focusing on the use of RT-LAMP to detect SARS-CoV-2^10–13^. We set up a SARS-CoV-2 RT-LAMP assay using the WarmStart Colorimetric LAMP 2X Mastermix commercialised by New England Biolabs (NEB), which allows for visual assessment of DNA amplification. Alternatively, DNA amplification can be quantified on a real-time PCR machine by complexing the reaction with the DNA dye SYTO 9^10^. We made use of primers developed by Zhang *et al*. targeting the nucleocapsid phosphoprotein (N gene) of SARS-CoV-2^10^ and, in a parallel reaction, primers that detect human 18S rRNA to control for specimen quality^9^.

Our results demonstrate that within the CCC pipeline, RT-LAMP can readily replace RT-qPCR as a means for detecting SARS-CoV-2 transcripts within RNA extracted from nose-throat swabs and endotracheal secretions/bronchoalveolar lavage fluid. RT-LAMP for the N gene shows absence of non-specific amplification and cross-reactivity with other human coronaviruses or respiratory viruses, and displays a sensitivity threshold almost equivalent to the gold standard RT-qPCR. Switching to RT-LAMP translates into a ten-fold decrease in total reagent cost and a potential four-fold increase in pipeline output. Additionally, we provide preliminary data suggesting that RT-LAMP can be performed without prior RNA extraction, allowing rapid and cost-effective testing that could potentially be extended to point-of-care. The entire workflow was validated under extended governance by public health authorities during the pandemic and inspected by a qualified UKAS assessor against GenQA guidelines to verify compliance to ISO 15189:2012 equivalent standard (US equiv. CAP/CLIA). As such, the standard operating procedure (SOP) developed here is ready to be deployed for diagnostic testing of SARS-CoV-2.

## Material and Methods

### CCC Pipeline Procedures and HSL RT-qPCR

All the samples processing through the CCC pipeline was done in accordance with SOPs described recently^5^. A redefined reaction, running, and reporting SOP can be found in the Supplemental Methods.

### RT-LAMP Assay

RT-LAMP reaction was performed in a total volume of 15 µL, mixing 7.5 µL WarmStart Colorimetric LAMP 2X Master Mix (New England Biolabs, M1800), 1.5 µL of 10X primer mix, 1.5 µL SYTO 9 Green Fluorescent Nucleic Acid Stain (ThermoFisher Scientific, S34854), 1.5 µL of Nuclease-free water (Ambion, AM9932) and 3 µL of sample unless stated otherwise. 10X SYTO 9 solution at 5 µM in nuclease free water was prepared for a final concentration of 0.5 µM in the final RT-LAMP reaction. 10X primer mix was prepared from 100 µM desalted DNA primers obtained from Sigma Custom Oligos Service. 10X primer mixes for N gene and 18S contained FIP and BIP primers at 16 µM, F3 and B3 at 2 µM, LF and LB at 4 µM. RT-LAMP was ran following a Standard Operating Procedure (cf. Appendix) on a 7500, 7500 Fast, QuantStudio 3, 5, or 7 Real-Time PCR System (Applied Biosystems). A negative no template control (NTC) and a positive control (supplied by HSL, SARS-CoV-2 clinical sample) were included on every run. Experiments utilising laboratory grown SARS-CoV-2 were performed in containment level 3 at the Francis Crick Institute (FCI) by trained personnel according to health and safety guidelines.

We found that the assembled reaction mix was unstable when kept at 4°C for more than a few hours, and sensitive to freeze-thaw when kept at −20°C for more than a day. The RT-LAMP should be performed using freshly prepared reaction mix. The individual components should be stored at −20°C until use and avoid repeated freeze/thaw cycles.

### Accreditation and Governance

As outlined recently^5^, the CCC was formed in partnership with HSL, a UCLH UKAS accredited lab, who already had a COVID-19 RT-qPCR test in scope. All samples were received and communicated by HSL under their accreditation and the CCC RT-LAMP assay was validated against their existing RT-qPCR test and the CCC’s validated RT-qPCR test, which uses a CE marked commercial kit (BGI). Given the urgent timeframe required to implement testing, it was not possible to secure official clinical laboratory accreditation for the Francis Crick Institute. However, full measures were taken to ensure that the CCC RT-LAMP test was evaluated, verified and performed for diagnostic use in an environment that adhered to equivalent international standards (ISO 15189:2012, US equiv. CAP/CLIA), overseen and audited by HSL. These measures were implemented under the advice and oversight of registered professionals from existing nearby ISO accredited medical laboratories, and included writing and following clinical diagnostic SOPs for every stage of the pipeline from sample reception, processing to result reporting by qualified clinical scientists prior to results being communicated to patients by HSL. Additional SOPs were followed for sample storage, disposal of materials, batch certification of reagents and incident reporting. Appropriate risk assessments, training and competency assessment procedures were established and documented. Record sheets were created to document the receipt, batch acceptance testing, and start/end of use dates for key reagents and consumables. An inventory of all key equipment was compiled which, where appropriate, included details of service and calibration records. Systems were also established for the control of all key documents (version implementation, distribution and acknowledgement), audit trailing (what samples were tested when, by whom, with what equipment and using which consumable/reagent batches), and the recording of all untoward incidents/issues (thus facilitating appropriate investigation, rectification and recurrence prevention). Samples were barcoded and tracked using the Crick Clarity library information management system (LIMS). All key documents are available at https://www.crick.ac.uk/research/covid-19/covid19-consortium. NHS governance was extended by a specific memorandum of understanding for diagnostic testing between UCLH and the Francis Crick Institute enabled by NHS England. Assurance of the pipeline was performed in collaboration with GenQA, following their checklist for non-accredited laboratories and the lab and CCC workflow were inspected by a qualified UKAS assessor against the GenQA guidelines to verify compliance to IS015189:2012 equivalent standard.

## Results

### RT-LAMP test validity

RT-LAMP was performed with 3 µL of RNA in a total reaction volume of 15 µL unless stated otherwise. Each sample was tested in two separate reactions, one with primers targeting the N gene of SARS-CoV-2, and the second targeting human 18S rRNA to control for specimen integrity and quality (Figure 1A). The RT-LAMP mastermix contains a colorimetric pH indicator that turns from pink to yellow upon DNA amplification (Figure 1B). In addition, we benchmarked the RT-LAMP method by measuring DNA amplification using a SYBR based dye in a real-time PCR machine. When measuring fluorescence every minute (‘cycle’), double stranded DNA accumulation follows a characteristic exponential amplification phase that eventually plateaus (Figure 1C and 1D).

**Figure 1.**
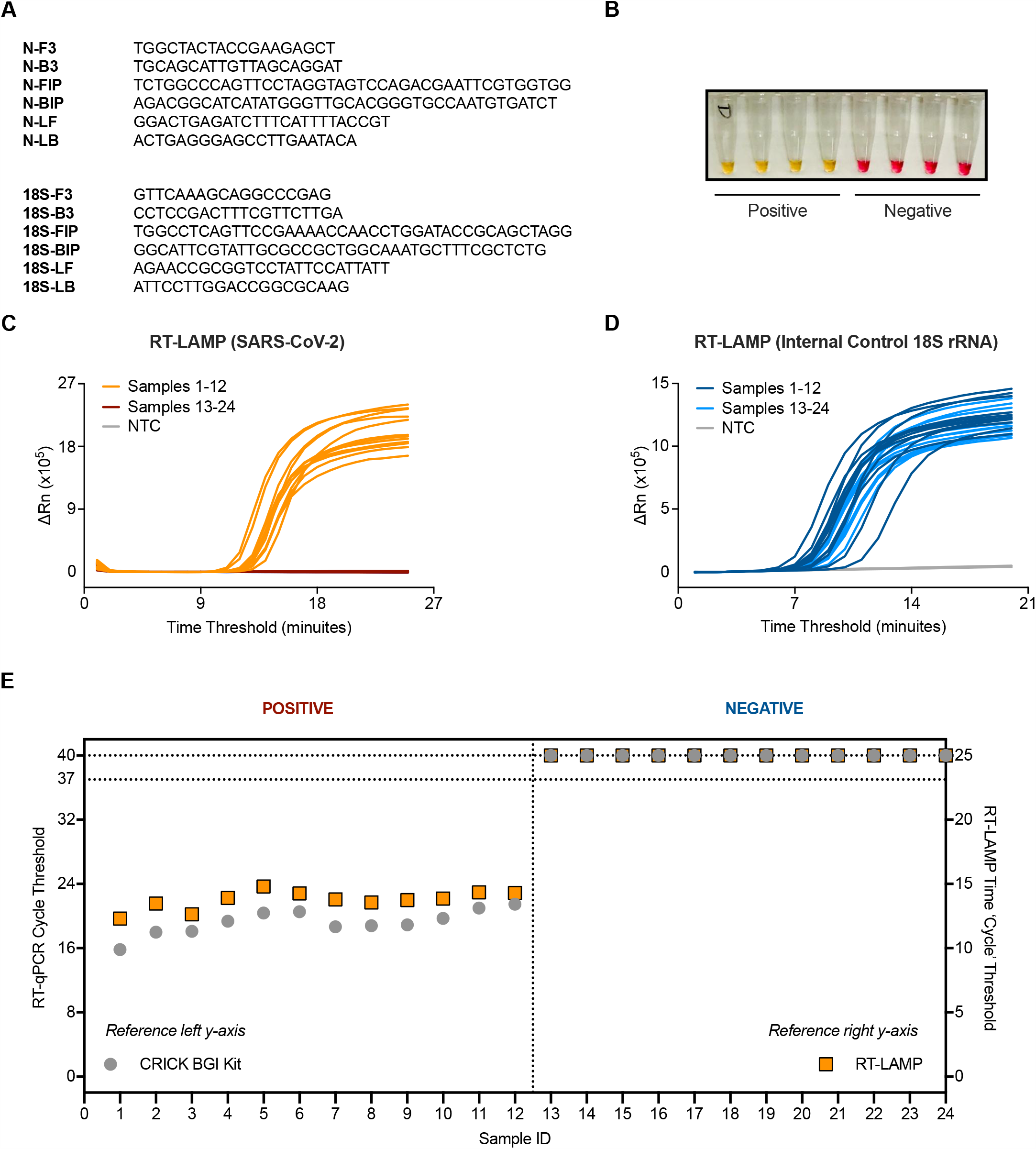
Validation of SARS-CoV-2 detection by RT-LAMP targeting the N gene. (**A**) The N gene of SARS-CoV-2 was targeted using primers designed by Zhang *et al*.^10^, whilst the internal control implemented utilised primers for 18S rRNA initially published by Lamb *et al*.^9^. (**B**) Upon DNA amplification, a colorimetric pH dye in the reaction mix will change from pink to yellow if the target is present. Four SARS-CoV-2 positive and four SARS-CoV-2 negative patient samples are shown. (**C-D**) Amplification curves from 24 patient samples assessed by SARS-CoV-2 (N gene) RT-LAMP (**C**) or 18S (**D**) in a Real-Time PCR machine. (**E**) Dot plot of the SARS-CoV-2 Ct values from (**C-D**) compared to Ct values derived from the Crick’s RT-qPCR diagnostic pipeline using the CE marked BGI kit. Ct values of ‘undetermined’ are plotted as “Ct = 40” (left y-axis) or “Ct = 25” (right y-axis) for illustrative purposes. Data are normalised by assay run/cycles determined by the two methods for comparative purposes. The clinical call from the reference laboratory is indicated above the graph.

An initial characterization of the technique was performed using 24 RNA samples purified from patient nasopharyngeal swabs, 12 of which were positive for SARS-CoV-2 as assessed by the CCC pipeline via RT-qPCR^5^. RT-LAMP could detect SARS-CoV-2 in the 12 positive samples with no amplification detected in all 12 negative samples, displaying 100% concordance to our current clinical diagnostic platform (Figures 1C and 1E). Internal control signal was detected in all samples (Figure 1D).

### RT-LAMP background noise and specificity

The background signal of the SARS-CoV-2 RT-LAMP assay was assessed by running RNA elution buffer (7 samples) or nuclease free water (7 samples) no template controls (NTCs) performed 4 separate times by 4 different operators. Non-specific signal could be detected in one well of one experiment after 25 minutes (Figure 2A). A similar experiment was performed for the 18S rRNA internal control RT-LAMP assay and revealed signal in NTCs after 20 minutes (Figure 2B). Based on these NTC data, a detection threshold was set for each RT-LAMP assay which eliminated all false positives in the N gene RT-LAMP assay and 21 of 23 false positives in the 18S rRNA RT-LAMP assay (Figure 2C). This translated to a maximum run time for each reaction: 25 minutes for N gene and 20 minutes for 18S (Figure 1C). Implementing these criteria in a separate experiment yielded no false positives from 47 NTCs in the SARS-CoV-2 RT-LAMP protocol and 4.5% false positives in the 18S rRNA RT-LAMP assay (all with a time threshold > 19) (Figure 2D-E). We then asked if melting curves obtained from thermocycler runs could be used to help discriminate true positives from false positives. N gene and 18S rRNA RT-LAMP reactions each gave single peaks in melt curve plots (Figure S1A) and consistent melting temperature values (Figure S1B) over multiple experiments performed on positive and negative patient samples. Moreover, positive samples that amplified near the assay endpoint (*i*.*e*., high time threshold) still displayed a clear melting curve peak consistent with being true positives (Figure S2A) and were distinguishable from false positives (Figure S2B). Therefore, when implementing the assay using a real-time PCR machine (thermocycler) with SYBR-based detection, incorporation of a traditional melting curve stage allows for elimination of false positives and increases reporting confidence for SARS-CoV-2 positive samples (see Supplemental Methods).

**Figure 2.**
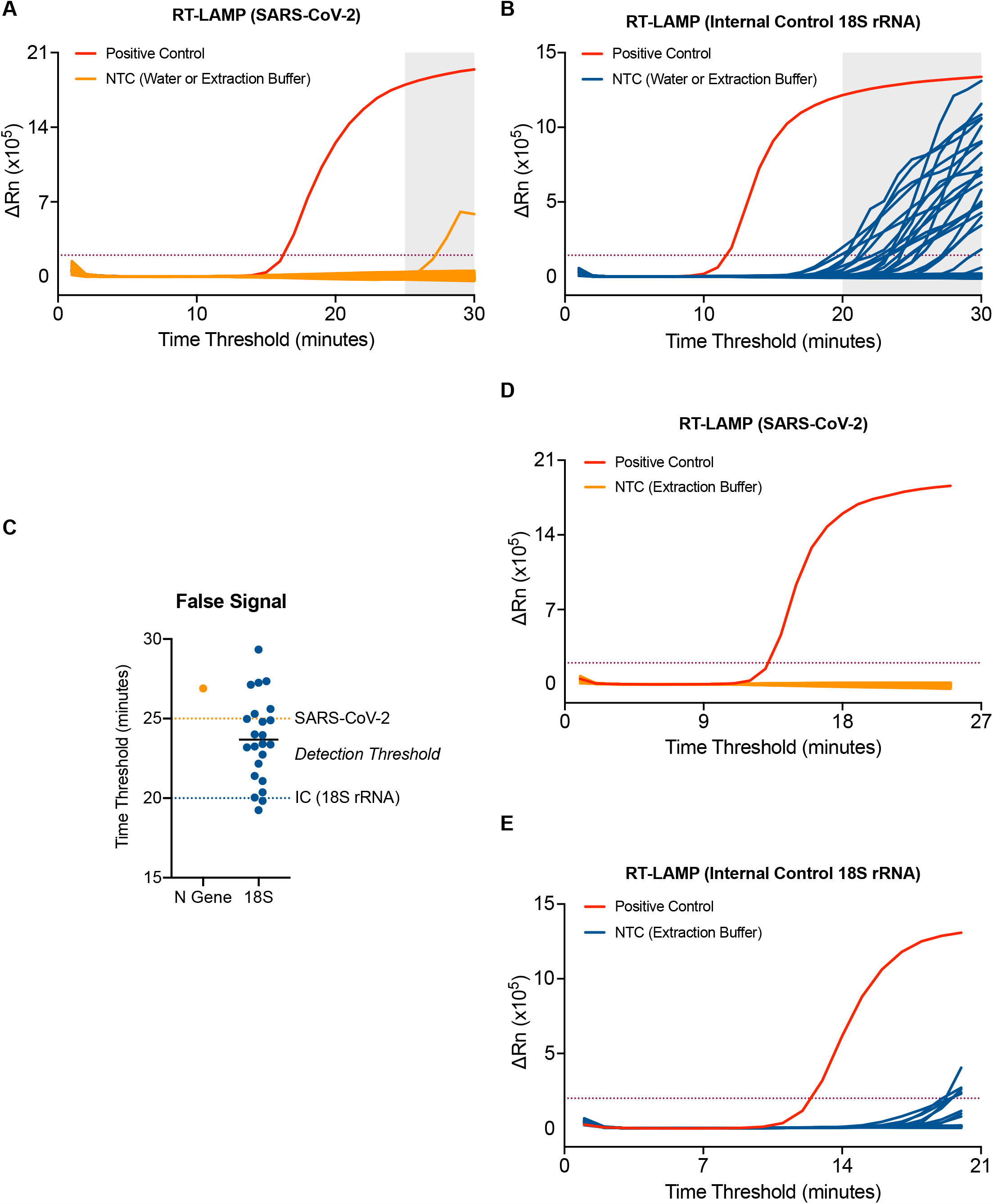
Optimisation of the RT-LAMP assay for accurate detection. (**A-B**) 7 samples of water and RNA elution buffer from the CCC pipeline were run 4 independent times by 4 distinct operators by SARS-CoV-2 (**A**) or 18S (**B**) RT-LAMP. (**C**) Dot plot of ‘Ct’ values assessed in (**A-B**). Assay endpoint detection threshold was set for 18S (dashed blue line) and SARS-CoV-2 (dashed orange line) based on these data. (**D-E**) RT-LAMP amplification data from 47 samples of RNA elution buffer using the newly established assay endpoint of 25 min for SARS-CoV-2 (**D**) and 20 minutes for 18S (**E**).

To further assess the specificity of the N gene RT-LAMP assay we first determined possible cross-reactivity with human RNA. 95 wells of RNA from a human cell line extracted by the CCC pipeline gave no signal in the assay (Figure 3A). To assess the assay’s specificity for SARS-CoV-2, the RT-LAMP assay was performed on RNA purified from clinical samples from COVID-19 negative patients infected with a variety of RNA viruses, including seasonal coronaviruses strains 229E, NL63, OC43 and HKU1, MERS-CoV, Influenza A and B viruses, metapneumovirus (MPV), respiratory syncytial virus (RSV) or parainfluenza viruses type 3 and 4 (PIV3, PIV4). All samples gave a signal in the 18S rRNA RT-LAMP, confirming the presence of human RNA (Figure 3B). With the N gene RT-LAMP, no signal was observed with any of the 14 distinct virus containing specimens tested, confirming that the assay is highly specific for SARS-CoV-2 (Figure 3B).

**Figure 3.**
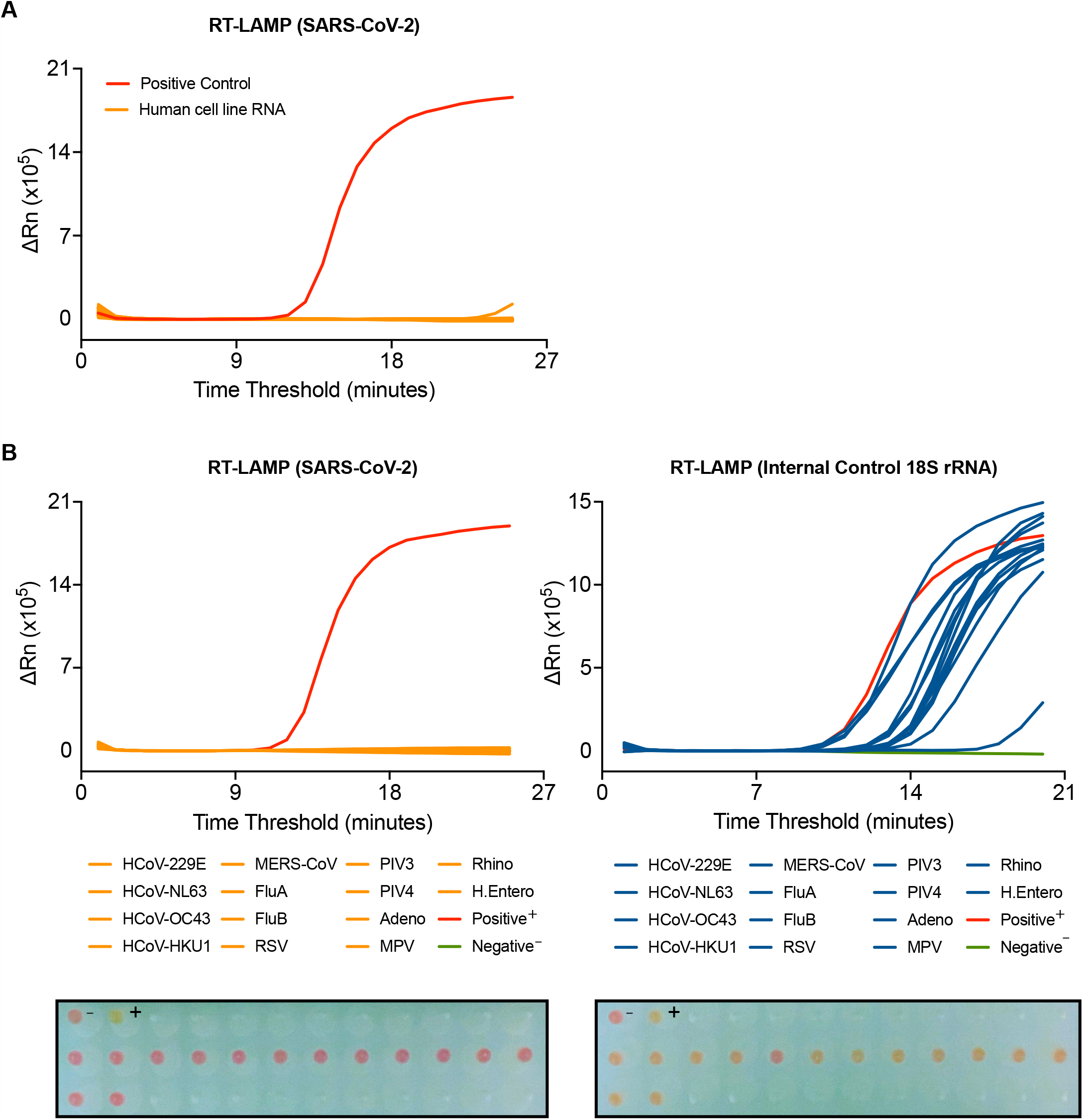
SARS-CoV-2 RT-LAMP is highly specific. (**A**) SARS-CoV-2 RT-LAMP was performed on 95 wells of human cell line RNA extracted by the CCC pipeline. (**B-C**) RT-LAMP targeting SARS-CoV-2 (**B**) or 18S (**C**) was performed on COVID-19 negative patient samples with positively identified with other viral infections, including human coronaviruses (HCoV), influenzas (Flu), respiratory syncytial virus (RSV), parainfluenzavirus (PIV), adenovirus (Adeno), metapneumovirus (MPV), rhinovirus (Rhino), and human enterovirus (H. Entero). The colorimetric read-out from the actual run is depicted below. NTC (-) and positive control (+).

### RT-LAMP sensitivity and precision

In order to ascertain the sensitivity of the N gene RT-LAMP assay, RNA from a SARS-CoV-2 quantified gene copy number standard obtained from the UK National Institute for Biological Standards and Control (NIBSC) was extracted and assessed by limiting dilution. The results indicate that the limit of reliable detection of the N gene RT-LAMP assay is between 500-1000 copies of N gene (Figure 4A-B). This was confirmed using 10-fold serial dilutions of RNA extracted from laboratory-grown SARS-CoV-2 quantified using the NIBSC standard (Figure 4C). Notably, a highly linear response was observed, even in the presence of 1% Triton-X 100, which has been proposed as a means of inactivating SARS-CoV-2^14^.

**Figure 4.**
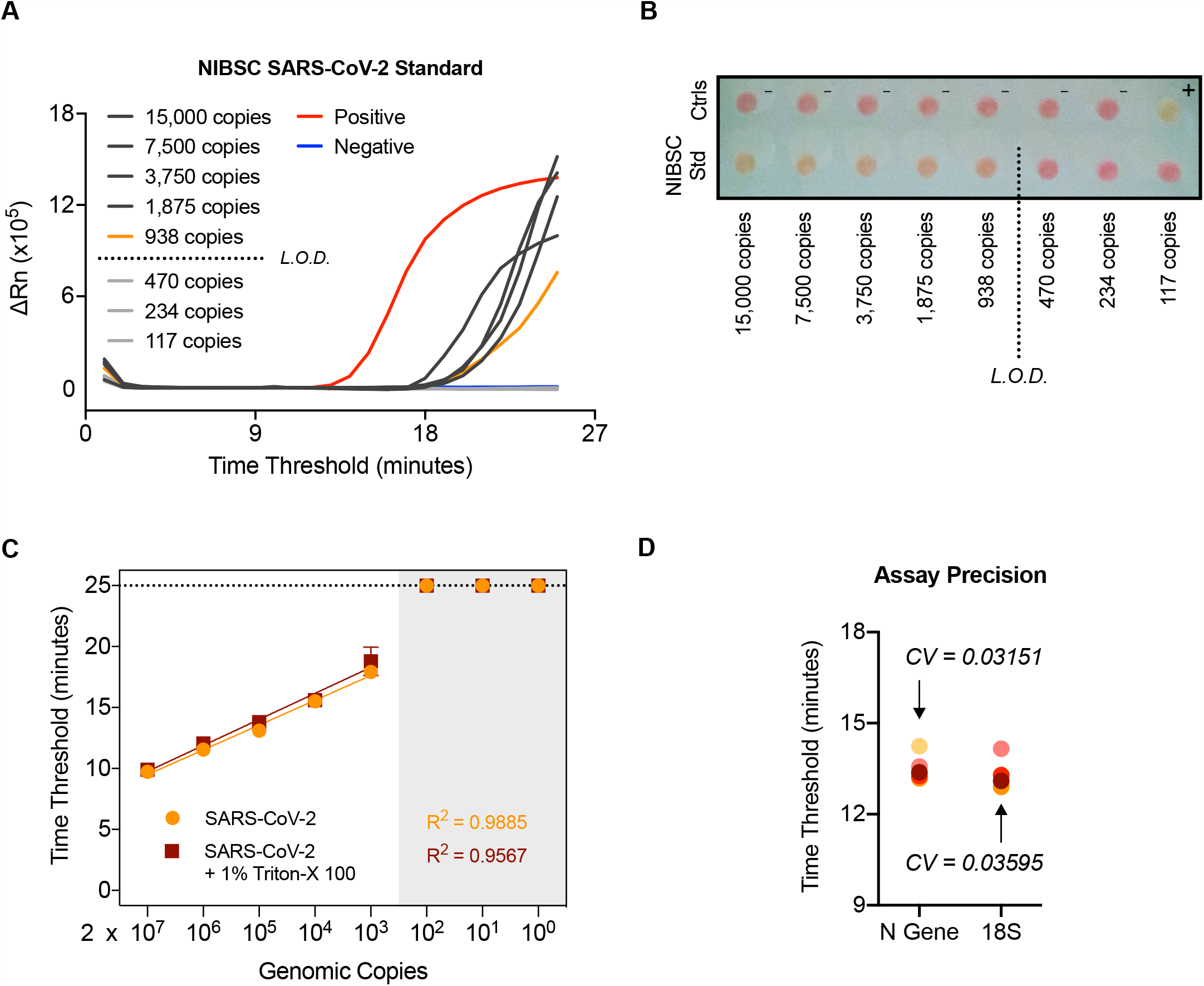
SARS-CoV-2 RT-LAMP is highly sensitive, robust, and precise. (**A-B**) NIBSC SARS-CoV-2 standard was serially diluted and the indicated number of copies was assessed by N gene RT-LAMP. Amplification curves shown with the limit of detection (L.O.D.) determined by the presence or absence of amplification following the depicted dilution in (**A**) or via colorimetric read-out (**B**). (**C**) RNA extracted from laboratory grown SARS-CoV-2 was serially diluted 10-fold and assessed by RT-LAMP in the presence or absence of 1% Triton-X 100. (**D**) A COVID-19 positive patient sample was RNA extracted independently 5 times through the CCC pipeline and subjected to 5 independent N gene RT-LAMP reactions. Assay precision for N gene and 18S was determined by calculating the coefficient of variation between Ct values observed (CV).

The RT-LAMP assay reproducibility and precision were determined by extracting RNA 5 times from a confirmed COVID-19 positive patient sample through the CCC pipeline and assessing by N gene and 18S RT-LAMP in 5 independent experiments, performed by two different operators (Figure 4D). The coefficient of variation was 0.03151 for the N gene RT-LAMP analysis and 0.03595 for the 18S rRNA internal control. We also verified that RT-LAMP can be performed in a 384-well plate format using a QuantStudio 5 real-time PCR machine with equivalent results (Figure S3).

### Clinical validation of RT-LAMP against two clinical diagnostic RT-qPCR assays

The RT-LAMP assay was benchmarked against the RT-qPCR methods used by the reference clinical diagnostic laboratory, HSL, and that used by the CCC validated clinical diagnostic platform by assessing 37 clinical samples processed in parallel by both laboratories with duplicate RT-qPCR analyses. RNA extracted by the CCC pipeline was then tested by RT-LAMP in duplicate runs. RT-LAMP detected 16 positives in both experiments, which was 100% concordant with results obtained by the CCC’s RT-qPCR assay using a CE marked kit from BGI and HSL’s N gene RT-qPCR assay. However, positives with Ct > 35 near the limit of detection (Ct = 37) of the BGI RT-qPCR assay, which were termed ‘borderline positives’, were not consistently detected by the N gene RT-LAMP assay (Figure 5A), despite those samples displaying clear signal for 18S rRNA amplification (data not shown). In an additional set of experiments, 71 clinical specimens in viral transport medium (VTM) stored at HSL were subjected to RNA extraction through the CCC pipeline and examined by HSL’s RT-qPCR and CCC’s RT-LAMP assay in duplicate (Figure 5B). Again, samples with Ct < 39 in the HSL RT-qPCR analysis (which runs for 45 cycles as opposed to 40 in the BGI assay) were confidently detected by RT-LAMP, whilst samples with Ct values near the limit of detection (39 < Ct < 41), indicating very low levels of SARS-CoV-2 RNA, displayed inconsistent amplification by N gene RT-LAMP despite showing signal for 18S rRNA RT-LAMP amplification (data not shown). Altogether, these data demonstrate that RT-LAMP performed on extracted RNA accurately detects SARS-CoV-2 in clinical samples as verified by a clinical diagnostic laboratory and a validated clinical diagnostic pipeline. However, samples with Ct values within two cycles of the limit of detection of either RT-qPCR assays, were not consistently identified in duplicate RT-LAMP runs, with detection for some samples in one run, but not always in the other. These results suggest that RT-LAMP is slightly less sensitive than RT-qPCR when performed with extracted RNA. However, it is important to bear in mind that the RT-LAMP assay was performed during these validation experiments with a third of the input RNA used by either RT-qPCR methods during these validation experiments (3 µL versus 10 µL), partly due to limitations in sample availability. Although not examined here, recent studies have proposed that the addition of guanidine containing compounds in millimolar amounts can improve the detection sensitivity of the RT-LAMP assay^11,15^.

**Figure 5.**
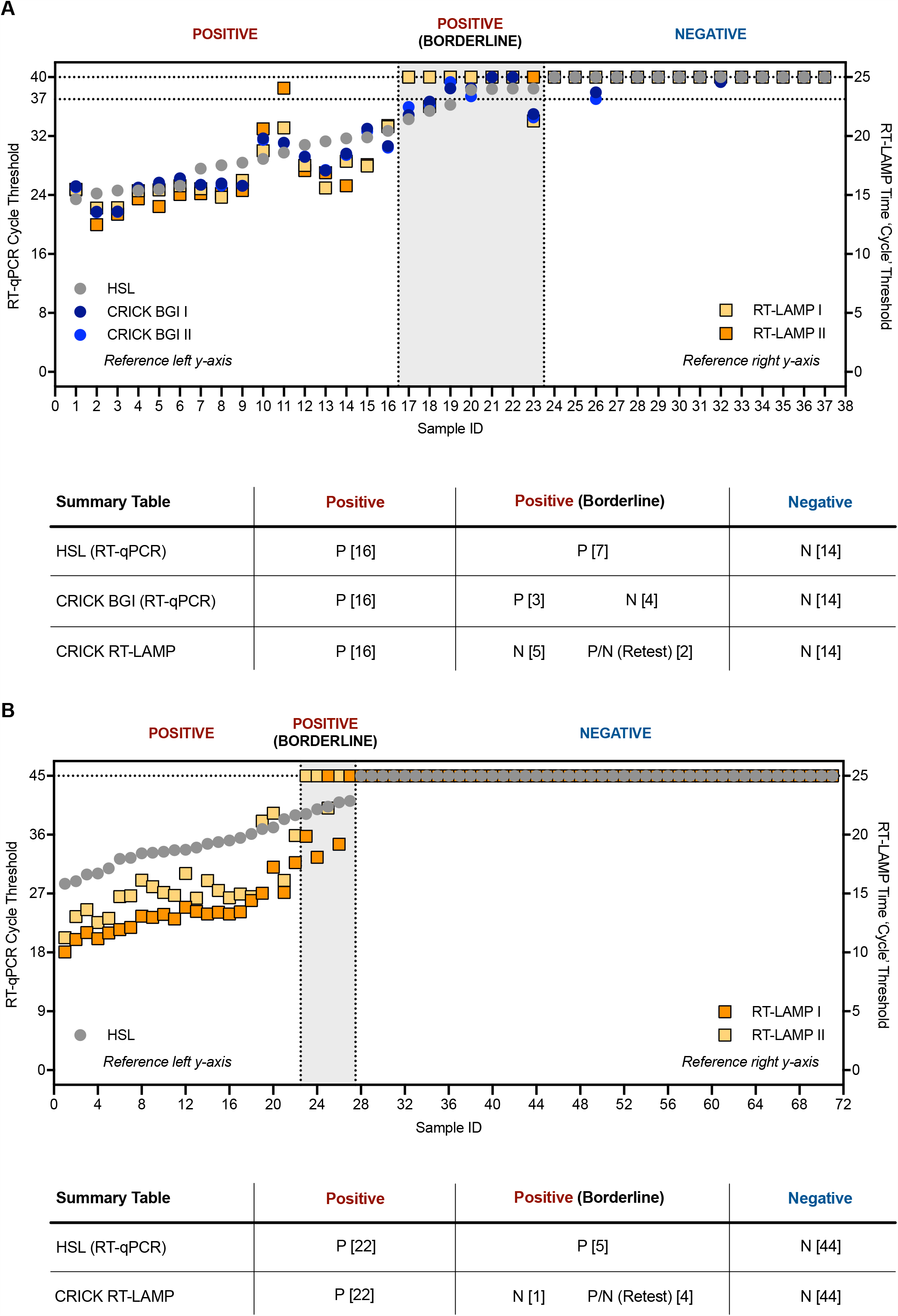
Clinical validation of SARS-CoV-2 RT-LAMP. (**A**) 37 patient samples were processed in parallel by HSL and the CCC pipeline and interrogated by HSL’s RT-qPCR (N gene) and the CCC’s BGI RT-qPCR (ORF1a) in duplicate. RNA left from the CCC pipeline was assessed in two separate experiments by N gene RT-LAMP. The graph indicates Ct values for HSL and CCC RT-qPCR runs by left y-axis and ‘Ct’ time thresholds for RT-LAMP via the right y-axis. Data are normalised by assay run/cycles determined by the two methods for comparative purposes. The clinical call from the reference laboratory is depicted above the summary table provided below. Positives (P) with low Ct values were reliably detected by RT-LAMP, whilst ‘borderline positive’ (samples with Ct values near the limit of detection of each RT-qPCR assay) were inconsistently detected, displaying no visible amplification at times – negative (N). (**B**) 71 VTM samples from HSL were RNA-extracted through the CCC pipeline and interrogated by HSL RT-qPCR (45 cycles), or by two independent N gene RT-LAMP experiments (25 minutes/cycles).

### Bypassing RNA extraction

Lastly, we asked if RT-LAMP could be performed on dry swabs without the need for RNA extraction. Recent work shows that 0.5% Triton X-100 inactivates SARS-CoV-2^14^, and we observed that performing RT-LAMP with samples diluted in 1% Triton X-100 does not affect the sensitivity of the assay (Figure 4C). We reverse-engineered swab specimens by coating hospital-grade swabs in serial dilutions with HEK293T cell supernatant of SARS-CoV-2 crude suspension generated in the laboratory. Swabs were left to dry overnight, and placed in 0.5% Triton X-100 for 15-30 minutes at room temperature before assessment. The RT-LAMP test used 4.5 µL of the solubilised material to maximise the sensitivity of the assay (Figure 6). In parallel, samples were processed by the CCC pipeline and interrogated by RT-qPCR with a calculated equivalent amount of RNA. ‘RT-LAMP Pre’ (without RNA purification) results were 94% concordant to those generated by the CCC pipeline using RT-qPCR, suggesting that RT-LAMP may potentially be performed without traditional RNA extraction. However, the use of RT-LAMP on material from swabs without RNA extraction requires clinical validation using *bona fide* nasopharyngeal specimens to ascertain reliability and compatibility with various transport media.

**Figure 6.**
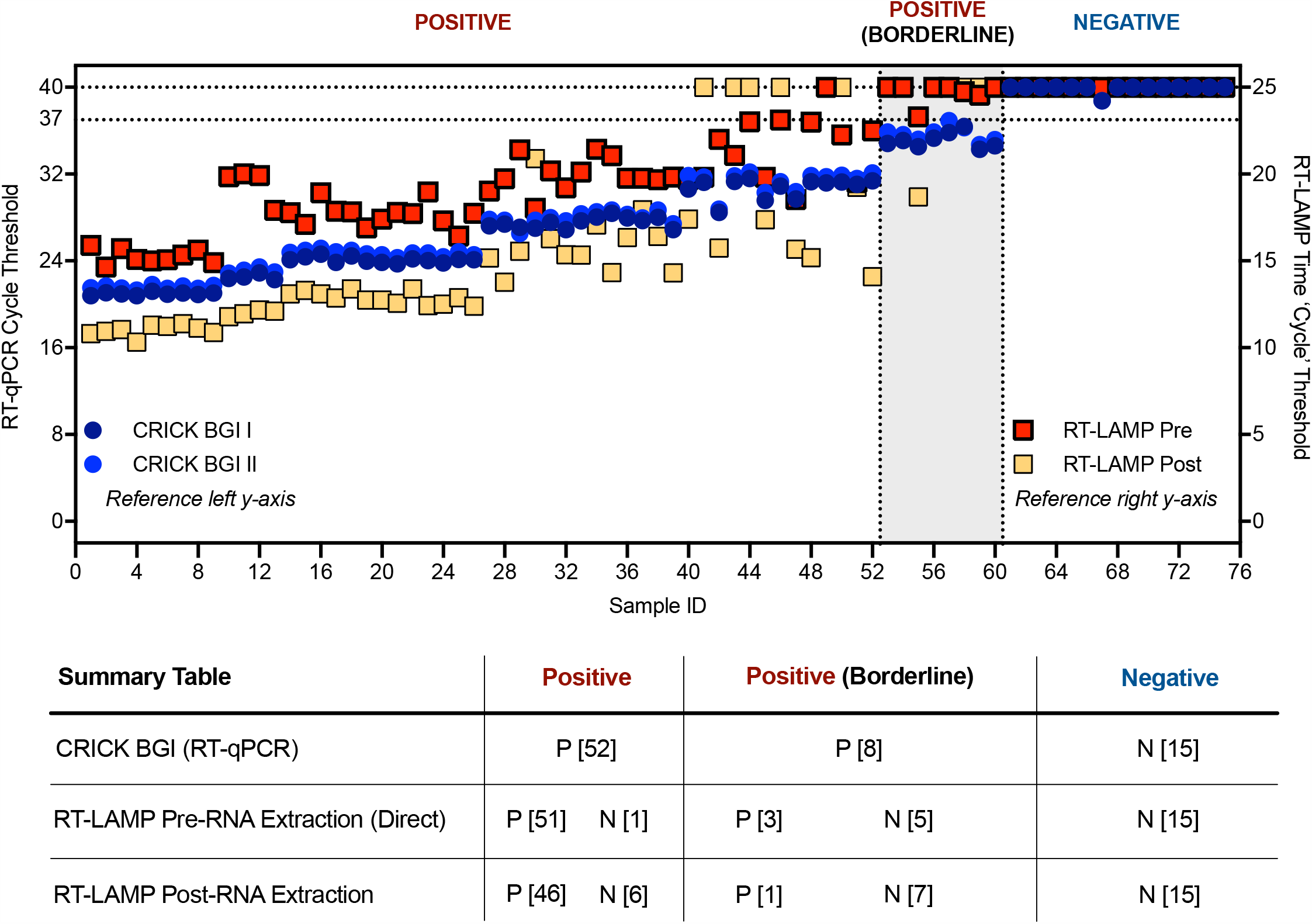
Comparison of SARS-CoV-2 RT-LAMP on direct versus RNA extracted mock swabs. 75 mock clinical grade swabs were generated by dipping into either SARS-CoV-2 virus suspension in a limiting dilution series with supernatant derived from human cell line (293T) culture supernatant or human cell culture supernatant alone (negative) and dried overnight. The following day, the swabs were suspended in 0.5% Triton-X 100 in water for 15-30 min at room temperature, a regimen recently shown to inactivate SARS-CoV-2^14^, and 4.5 µL of the sample was assessed by RT-LAMP assay (RT-LAMP Pre). In parallel, an equivalent genomic eluent was subjected to standard viral inactivation and RNA extraction by the CCC pipeline and also assessed by RT-qPCR in duplicate. The resulting RNA was also assessed by RT-LAMP (RT-LAMP Post). Dot plot demonstrating the SARS-CoV-2 values obtained from the 75 in-house generated samples tested by RT-LAMP assay directly from dry swabs without traditional RNA extraction procedures (red squares) compared to Ct values derived from the CCC RT-qPCR BGI assay (dark and light blue dots) and the RT-LAMP assay following RNA extraction (yellow squares). The clinical call from the reference laboratory is indicated above the boxes. Ct values of ‘undetermined’ are plotted as “Ct = 40/25” for illustrative purposes.

## Discussion

Rapid and reliable detection of SARS-CoV-2 is required to efficiently diagnose infected individuals and to provide governments and health systems with guidance for treatment and quarantine strategies to reduce the risk of transmission. The CCC was formed to address a critical testing shortage in the London area, especially for frontline healthcare workers, with the additional goal of rapidly validating and disseminating SOPs for others to scale up their own diagnostic programs. The majority of testing is currently performed by RT-qPCR amplification of viral RNA obtained from nasopharyngeal samples. In the face of continued global demand and competition for reagents and resources and to minimise reliance on any singular testing strategy, we have outlined here procedures to utilise RT-LAMP as a cost-effective and high throughput alternative to RT-qPCR for detecting SARS-CoV-2 in clinical specimens.

Implementing this testing method would reduce the CCC’s operational costs ten-fold when accounting for all consumables (data not shown), provide the ability to scale up our output further (Figure S3), and decrease reporting turnaround time in comparison to our current method. We confirmed that the assay is highly specific for SARS-CoV-2 and displays lack of cross-reactivity with other respiratory viruses, including seasonal coronaviruses. Assay accuracy and robustness is denoted by an absence of false positives and the further introduction of a melt curve stage allows increased confidence in the identification and reporting of genuine positives. When performed with extracted RNA, the N gene RT-LAMP assay displays a sensitivity comparable to clinically approved RT-qPCR methods. Additionally, preliminary experiments demonstrate that RT-LAMP can be performed directly without RNA extraction, by inactivating virus-containing dry swabs in a detergent-based solution. However, these experiments did not make use of true nasopharyngeal swabs that are likely to contain material that can cause RNA degradation or inhibit the assay (unpublished observations).

Our methodology makes use of a real-time qPCR machine that measures DNA amplification using a SYBR based dye, allowing for real-time detection and standardised reporting. We did not test extensively if RT-LAMP could be assessed by the colorimetric indicator alone, although some of our data suggests that the colour change readout is concordant with the SYTO 9 dye results (Figures 1, 3 and 4). If RT-LAMP were coupled with a colorimetric read-out and the need for RNA extraction could be obviated, the result would be a testing modality that tremendously reduces the cost and time for SARS-CoV-2 diagnostics and allow its application at point-of-care and in remote areas where sophisticated testing infrastructures currently do not exist.

## Data Availability

All key documents are available at https://www.crick.ac.uk/research/covid-19/covid19-consortium

## Acknowledgments

This work was supported by The Rosetrees Trust and The John Black Charitable Foundation, as well as the Francis Crick Institute, which receives its funding from the UK Research Medical Council (FC001169, FC001078), Cancer Research UK (FC001169, FC001078) and the Wellcome Trust (FC001169, FC001078). E. Nastouli is funded by MRC, NIHR, GSK and H2020. S. Gandhi has an MRC Senior Clinical Fellowship. E.Z. Poirier and M.D. Buck are supported by EMBO Long-Term Fellowships (ALTF 536-2108 and ALTF 1096-2018) and Marie Skłodowska-Curie Individual Fellowships (832511 and 837951).

The authors wish to thank all of the CCC members and CAPTURE study members that have helped contribute reagents, advice, and time to this work.

**The Crick COVID-19 Consortium (CCC)**

Jim Aitken^1^, Zoe Allen^2^, Rachel Ambler^1^, Karen Ambrose^1^, Emma Ashton^2^, Alida Avola^1^, Samutheswari Balakrishnan^1^, Caitlin Barns-Jenkins^1^, Genevieve Barr^1^, Sam Barrell^1^, Souradeep Basu^1^, Rupert Beale^1,3^, Clare Beesley^2^, Nisha Bhardwaj^1^, Shahnaz Bibi^2^, Ganka Bineva-Todd^1^, Dhruva Biswas^3^, Michael J. Blackman^1,4^, Dominique Bonnet^1^, Faye Bowker^1^, Malgorzata Broncel^1^, Claire Brooks^2^, Michael D. Buck^1^, Andrew Buckton^2^, Timothy Budd^1^, Alana Burrell^1^, Louise Busby^2^, Claudio Bussi^1^, Simon Butterworth^1^, Fiona Byrne^1^, Richard Byrne^1^, Simon Caidan^1^, Joanna Campbell^5^, Johnathan Canton^1^, Ana Cardoso^1^, Nick Carter^1^, Luiz Carvalho^1^, Raffaella Carzaniga^1^, Natalie Chandler^2^, Qu Chen^1^, Peter Cherepanov^1^, Laura Churchward^6^, Graham Clark^1^, Bobbi Clayton^1^, Clementina Cobolli Gigli^1^, Zena Collins^1^, Sally Cottrell^2^, Margaret Crawford^1^, Laura Cubitt^1^, Tom Cullup^2^, Heledd Davies^1^, Patrick Davis^1^, Dara Davison^1^, Vicky Dearing^1^, Solene Debaisieux^1^, Monica Diaz-Romero^1^, Alison Dibbs^1^, Jessica Diring^1^, Paul C. Driscoll^1^, Annalisa D’Avola^1^, Christopher Earl^1^, Amelia Edwards^1^, Chris Ekin^7^, Dimitrios Evangelopoulos^1,3^, Rupert Faraway^1,3^, Antony Fearns^1^, Aaron Ferron^1^, Efthymios Fidanis^1^ Dan Fitz^1^, James Fleming^1^, Bruno Frederico^1^, Alessandra Gaiba^1^, Anthony Gait^2^, Steve Gamblin^1^, Sonia Gandhi^1,3,6^, Liam Gaul^1^, Helen M. Golding^1^, Jacki Goldman^1^, Robert Goldstone^1^, Belen Gomez Dominguez^2^, Hui Gong^1^, Paul R. Grant ^7^, Maria Greco^1^, Mariana Grobler^2^, Anabel Guedan^1^, Maximiliano G. Gutierrez^1^, Fiona Hackett^1^, Ross Hall^1^, Steinar Halldorsson^1^, Suzanne Harris^1^, Sugera Hashim^2^, Lyn Healy^1^, Judith Heaney^6^, Susanne Herbst^1^, Graeme Hewitt^1^, Theresa Higgins^1^, Steve Hindmarsh^1^, Rajnika Hirani^1^, Joshua Hope^1^, Elizabeth Horton^1^, Beth Hoskins^2^, Catherine F. Houlihan^6^, Michael Howell^1^, Louise Howitt^1^, Jacqueline Hoyle^2^, Mint R. Htun^1^, Michael Hubank^8,9^, Hector Huerga Encabo^1^, Deborah Hughes^8^, Jane Hughes^1^, Almaz Huseynova^1^, Ming-Shih Hwang^1^, Rachael Instrell^1^, Deborah Jackson^1^, Mariam Jamal-Hanjani ^3,6^, Lucy Jenkins^2^, Ming Jiang^1^, Mark Johnson^1^, Leigh Jones^1^, Nnennaya Kanu^3^, George Kassiotis^1^, Louise Kiely^2^, Anastacio King Spert Teixeira^1^, Stuart Kirk^7^, Svend Kjaer^1^, Ellen Knuepfer^1,12^, Nikita Komarov^1,3^, Paul Kotzampaltiris^5^, Konstantinos Kousis^1^, Tammy Krylova^1^, Ania Kucharska^1^, Robyn Labrum^5^, Catherine Lambe^1^, Michelle Lappin^1^, Stacey-Ann Lee^1^, Andrew Levett ^7^, Lisa Levett ^7^, Marcel Levi^6^, Hon Wing Liu^1^, Sam Loughlin^2^, Wei-Ting Lu^1^, James I. MacRae^1^, Akshay Madoo^1^, Julie A. Marczak^1^, Mimmi Martensson^1^, Thomas Martinez^1^, Bishara Marzook^1^, John Matthews^7^, Joachim M. Matz^1^, Samuel McCall^5^, Laura E. McCoy^3^, Fiona McKay^2^, Edel C. McNamara^1^, Carlos M. Minutti^1^, Gita Mistry^1^, Miriam Molina-Arcas^1^, Beatriz Montaner^1^, Kylie Montgomery^2^, Catherine Moore^10^, David Moore^3,6^, Anastasia Moraiti^1^, Lucia Moreira-Teixeira^1^, Joyita Mukherjee^1^, Cristina Naceur-Lombardelli^3^, Eleni Nastouli ^6,11^, Aileen Nelson^1^, Jerome Nicod^1^, Luke Nightingale^1^, Stephanie Nofal^1^, Paul Nurse^1^, Savita Nutan^2^, Caroline Oedekoven^1^, Anne O’Garra^1^, Jean D. O’Leary^1^, Jessica Olsen^1^, Olga O’Neill^1^, Nicola O’Reilly^1^, Paula Ordonez Suarez^1^, Neil Osborne^1^, Amar Pabari^7^, Aleksandra Pajak^1^, Venizelos Papayannopoulos^1^, Namita Patel^1^, Yogen Patel^5^, Oana Paun^1^, Nigel Peat^1^, Laura Peces-Barba Castano^1^, Ana Perez Caballero^2^, Jimena Perez-Lloret^1^, Magali S. Perrault^1^, Abigail Perrin^1^, Roy Poh^5^, Enzo Z. Poirier^1^, James M. Polke^5^, Marc Pollitt^1^, Lucia Prieto-Godino^1^, Alize Proust^1^, Clinda Puvirajasinghe^2^, Christophe Queval^1^, Vijaya Ramachandran^2^, Abhinay Ramaprasad^1^, Peter Ratcliffe^1^, Laura Reed^2^, Caetano Reis e Sousa^1^, Kayleigh Richardson^1^, Sophie Ridewood^1^, Fiona Roberts^1^, Rowenna Roberts^2^, Angela Rodgers^1^, Pablo Romero Clavijo^1^, Annachiara Rosa^1^, Alice Rossi^1^, Chloe Roustan^1^, Andrew Rowan^1^, Erik Sahai^1^, Aaron Sait^1^, Katarzyna Sala^1^, Theo Sanderson^1^, Pierre Santucci^1^, Fatima Sardar^1^, Adam Sateriale^1^, Jill A. Saunders^1^, Chelsea Sawyer^1^, Anja Schlott^1^, Edina Schweighoffer^1^, Sandra Segura-Bayona^1^, Rajvee Shah Punatar^1^, Joe Shaw^2^, Gee Yen Shin^6,7^, Mariana Silva Dos Santos^1^, Margaux Silvestre^1^, Matthew Singer^1^, Daniel M. Snell^1^, Ok-Ryul Song ^1^, Moira J. Spyer ^3^, Louisa Steel^2^, Amy Strange^1^, Adrienne E. Sullivan^1^, Charles Swanton^1,3,6^, Michele S.Y. Tan^1^, Zoe H. Tautz-Davis^1^, Effie Taylor^1^, Gunes Taylor^1^, Harriet B. Taylor^1^, Alison Taylor-Beadling^2^, Fernanda Teixeira Subtil^1^, Berta Terré Torras^1^, Patrick Toolan-Kerr^1,3^, Francesca Torelli^1^, Tea Toteva^1^, Moritz Treeck^1^, Hadija Trojer^13^, Ming-Han C. Tsai^1^, James M.A.Turner^1^, Melanie Turner ^7^, Jernej Ule^1,3^, Rachel Ulferts^1^, Sharon P. Vanloo^1,3^, Selvaraju Veeriah^3^, Subramanian Venkatesan^1^, Karen Vousden^1^, Andreas Wack^1^, Claire Walder^2^, Philip A. Walker^1^, Yiran Wang^1^, Sophia Ward^1,3^, Catharina Wenman^2^, Luke Williams^1^, Matthew J. Williams^1^, Wai Keong Wong^6^, Joshua Wright^1^, Mary Wu^1^, Lauren Wynne^1^, Zheng Xiang^1^, Melvyn Yap^1^, Julian A. Zagalak^1,3^, Davide Zecchin^1,11^ and Rachel Zillwood^1^

^1^The Francis Crick Institute, London NW1 1AT, UK

^2^Great Ormond Street Hospital for Sick Children NHS Foundation Trust, London WC1N 3JH, UK.

^3^University College London, London WC1E 6BT, UK

^4^London School of Hygiene & Tropical Medicine, London WC1E 7HT, UK

^5^National Hospital for Neurology and Neurosurgery, University College London Hospitals, NHS Foundation Trust, London WC1N 3BG, UK

^6^University College London Hospitals, NHS Foundation Trust, London NW1 2PG, UK

^7^Health Services Laboratories, London WC1H 9AX, UK

^8^The Institute of Cancer Research, London SW7 3RP, UK

^9^The Royal Marsden Hospital, Surrey SM2 5NG, UK

^10^Public Health Wales, Heath Park, Cardiff CF14 4XW, UK

^11^University College London GOS Institute of Child Health, London WC1N 1EH, UK ^12^Department of Pathobiology and Population Sciences, Royal Veterinary College, University of London, London NW1 0TU, UK

^13^Royal Free London NHS Foundation Trust, London NW3 2QG, UK

**CAPTURE study**

Nicky Browne^1^, Kim Edmonds^1^, Yasir Khan^1^, Sarah Sarker^1^, Ben Shum^1^, Tim Slattery^1^, & Samra Turajlic^1,2^

^1^The Royal Marsden Hospital, Surrey, SM2 5NG, UK

^2^The Francis Crick Institute, London NW1 1AT, UK

## Supplementary Figure Legends

**Figure S1.**
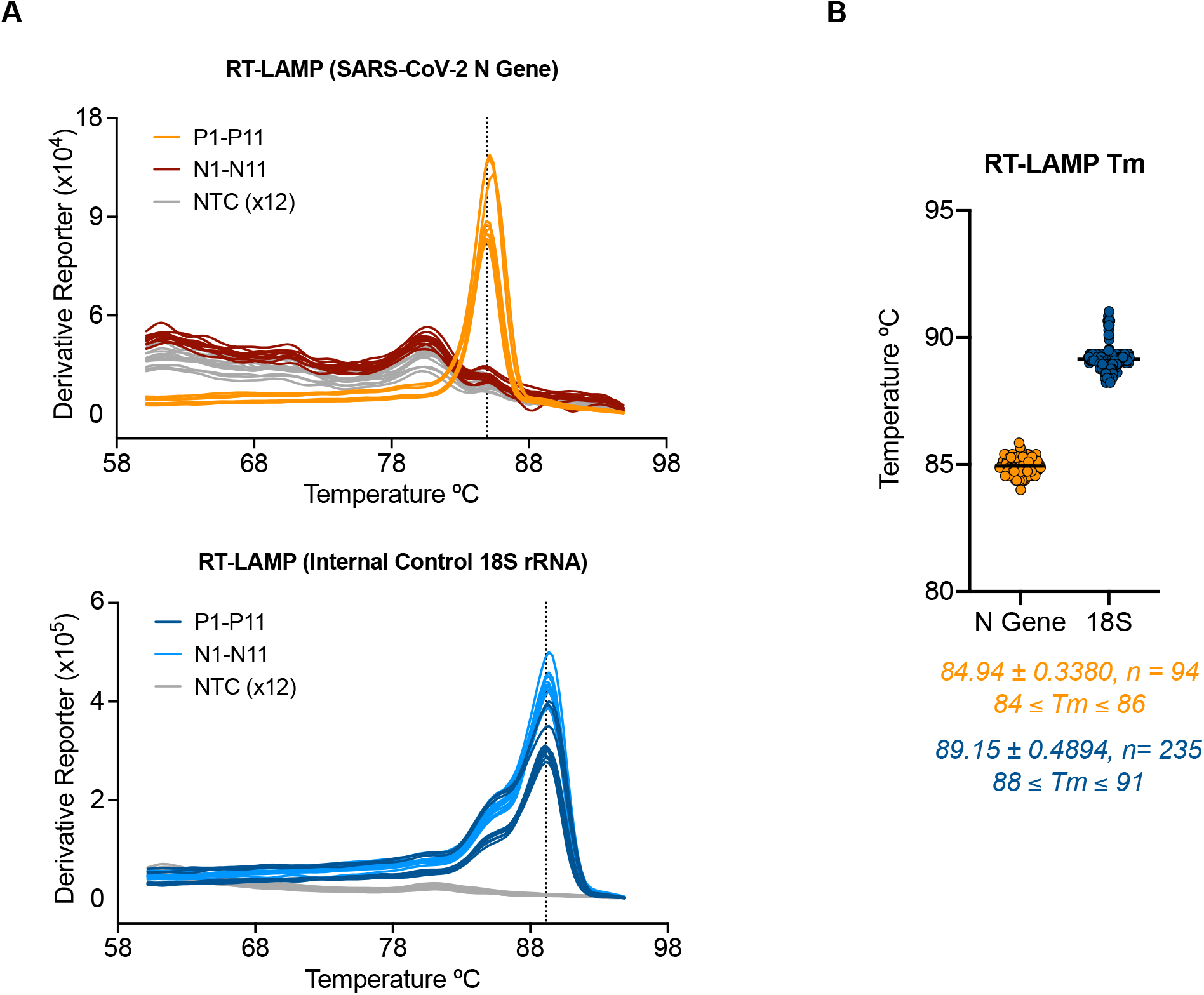
SARS-CoV-2 RT-LAMP and internal control 18S rRNA RT-LAMP display a consistent dissociation peak and melting temperature. (**A**) Dissociation curves are shown for 11 SARS-CoV-2 positive and 11 negative clinical samples along with 12 NTC wells tested by RT-LAMP N gene assay (top) and 18S assay (bottom). (**B**) Dot plot of the melting temperatures determined by 94 positive patient samples and 235 negative patient samples tested by RT-LAMP assay. Data are pooled from six separate experiments performed on 3 separate RT-PCR machines. The average melting temperature (Tm) is depicted by a dotted line in (**A**) and below the graph in (**B**) with the standard of deviation.

**Figure S2.**
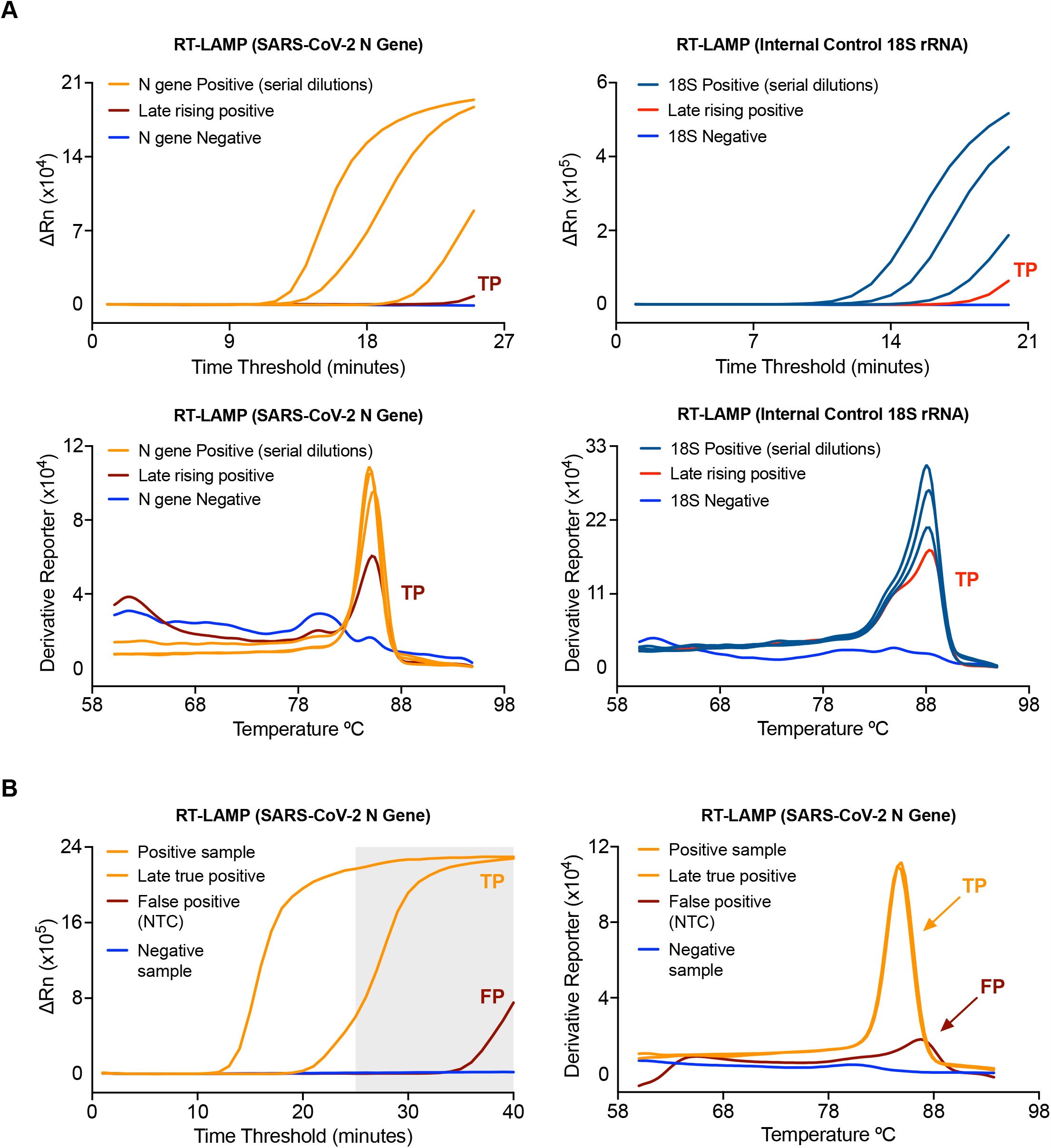
Inclusion of a melt curve stage complements the ability of RT-LAMP to accurately distinguish true SARS-CoV-2 positive samples. (**A**) Amplification curves (top) and dissociation curves (bottom) are depicted from a range of positive samples for each assay along with a negative sample for reference. TP indicates a true positive call based on amplification above background plus consistent melting curve and Tm consistent with genuine positives. (**B**) Amplification curve (left) and dissociation curve (right) are depicted of positive samples and NTCs. TP indicates the same criteria outlined in (**A**), whereas FP denotes a false positive, which despite amplifying above background, does not have a melting curve and temperature consistent with genuine positives.

**Figure S3.**
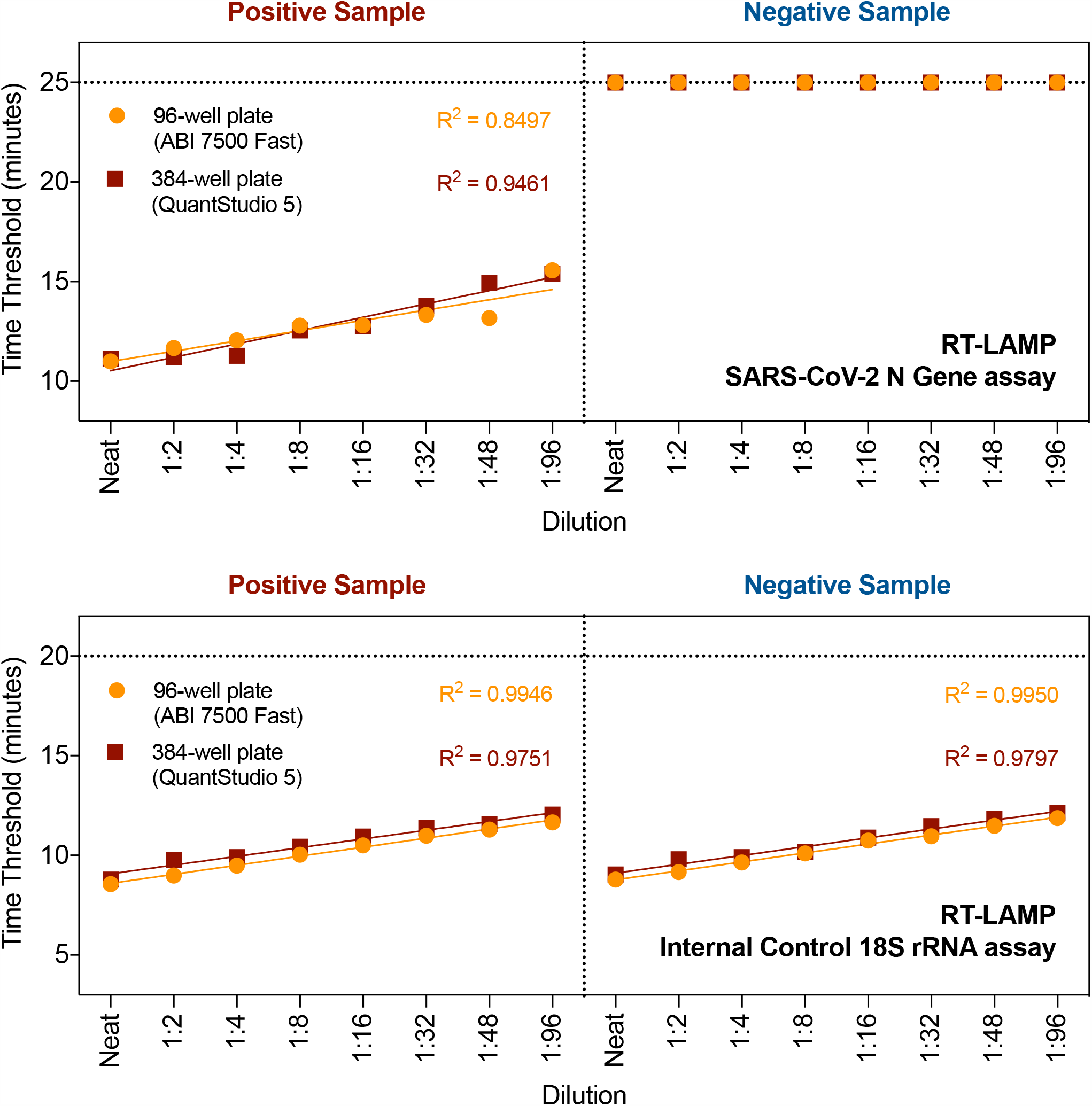
RT-LAMP yields identical results across different RT-PCR instruments and 96 versus 384-well plate formats. Dot plot comparing results from RT-LAMP assays (N gene, top; 18S, bottom) on the same clinical positive and negative sample serially diluted 2-fold using an ABI 7500 Fast or QuantStudio 5 machine (96-well vs. 384-well plate format).

